# Using an EMR to assess pediatric blood pressure: Challenges and opportunities in a nephrology cohort

**DOI:** 10.1101/2025.09.11.25335121

**Authors:** R Mah, A Tsampalieros, RJ Webster, I Terekhov, J Seymour, J Feber, RL Myette

## Abstract

**Background:** Hypertension is a prevalent condition in the pediatric population. Diagnosis and management can be challenging due to difficulties with accurate measurement techniques and complex diagnostic criteria. The widespread adoption of electronic medical records (EMRs) has revealed their potential for improving patient care and research. This study aims to assess the clinical utility of using EMR data to enhance the identification and evaluation of children with hypertension.

**Objectives:** The primary objectives of this research project were to utilize the EMR to extract anthropometric, demographic, and blood pressure-related data from patients seen in the nephrology clinic as well as describe and evaluate trends in hypertension assessment and treatment while also identifying areas for improvement.

**Design:** We performed a single center, retrospective cohort study using EMR data.

**Setting:** Children who had their initial visit at the nephrology clinic between January 1st, 2018, and January 1st, 2022, were included in the cohort.

**Methods:** Outpatients were identified using ICD-10 codes related to nephrology diseases. The EMR was reviewed to extract anthropometric, biochemical, and blood pressure data. A blood pressure (BP) index was calculated using systolic and diastolic BP values and the 2017 American Academy of Pediatrics (AAP) hypertension guidelines. The primary analysis categorized BP phenotypes. A secondary analysis using EMR and chart review, assessed whether elevated BP was appropriately managed, including scheduling follow-up visits, diagnosing white-coat hypertension, or initiating pharmacological or non-pharmacological interventions.

**Results:** A total of 1,469 children aged 1–18 (median age 9.8 years) were newly referred to the nephrology clinic with complete data for BP index calculation. Many children were initially diagnosed as hypertensive, but across multiple visits were normotensive. Despite being hypertensive across multiple visits, we observed that many children had missing data following EMR extraction (∼20%). Furthermore, despite meeting criteria at visit one for hypertension, many children did not have follow up visits (∼20-30%). We identified that those children presenting with isolated elevated diastolic blood pressure elevations were more likely to have fewer BP measurements and were less likely to have BP-related follow up, likely reflecting the perceived benign nature of this phenomenon.

**Limitations:** This study’s retrospective, non-randomized design limits generalizability.

**Conclusions:** This study underscores the challenges in studying pediatric hypertension using an EMR, particularly highlighting missing values and decreased measurements as problematic.

## Introduction

Hypertension (HTN) is an increasingly prevalent condition worldwide (1,2) and is a growing issue in the pediatric population (3,4). A recent meta-analysis estimated the global prevalence of elevated blood pressure (BP) and HTN among individuals aged 19 years as 9.7% and 4.0%, respectively (5). Data from the National Health and Nutrition Examination Survey (NHANES) showed a rising prevalence of elevated BP and HTN in children aged 8–17 over a 12-year period (6). The risk of elevated BP has increased by 27%, mostly attributed to an increase in obesity, waist circumference, and sodium intake (6,7).

HTN is a primary modifiable risk factor for cardiovascular disease and is often referred to as the “silent killer” due to its asymptomatic nature while progressively damaging multiple organ systems. Uncontrolled HTN increases the risk of early vascular aging, left ventricular hypertrophy, stroke, kidney injury, and metabolic syndrome (8). Longitudinal studies have demonstrated that elevated BP in childhood strongly correlates with an increased risk of HTN in adulthood, particularly when present in older children and adolescents (9–11). Certain pediatric populations are at higher risk for HTN. Children with chronic conditions, such as obesity (12,13), sleep-disordered breathing (14), chronic kidney disease, proteinuria (15–17), and prematurity (18,19), demonstrate a higher prevalence of HTN. Early identification of children with abnormal BP levels is critical to improving BP control, slowing disease progression, and reducing the long-term risk of cardiovascular disease and target organ damage (20–23). Despite its importance, the diagnosis and management of pediatric HTN present unique challenges. Proper BP measurement, complex diagnostic criteria, and hesitancy in initiating antihypertensive medication are obstacles (24).

Electronic medical records (EMRs) provide a digital platform for storing a patient’s clinical information, including administrative, diagnostic, and interventional data. Initially developed to enhance patient care and streamline clinical workflows, EMRs have become valuable tools for research and quality improvement initiatives due to their accessibility and ability to handle large datasets. However, challenges persist in using EMRs for pediatric research, including incomplete documentation, inconsistent data formats, lack of standardized platforms, and documentation biases (25–27). The primary objectives of this research project were to utilize the EMR to extract anthropometric, demographic, and blood pressure-related data from patients seen in the nephrology clinic as well as describe and evaluate trends in hypertension assessment and treatment while also identifying areas for improvement.

## Methods

This retrospective chart review study was conducted at the Children’s Hospital of Eastern Ontario (CHEO). Data were extracted by the CHEO Research Institute (CHEO RI) Data Warehouse and transferred to a secure CHEO RI Projects database by the Data Warehouse Clinical Data Analyst. Additional data were manually extracted through chart reviews. The study posed minimal risk to participants. The CHEO Research Ethics Board (REB) gave ethical approval for this work (REB#22/128X); however, the need for consent was waived given the retrospective nature of this study. Patients were eligible for inclusion if they were aged 1 to 18 years old and were seen at the nephrology clinic at CHEO between January 1st, 2018, and January 1st, 2022. Patients were excluded if clinical data such as age, sex, height or blood pressure measurements were missing, as these were essential for calculating the blood pressure index. Only patients who had elevated blood pressure at their first clinic visit, a follow-up visit within one year of the initial appointment, and another visit within the following year were included in the full analysis. Demographic, anthropometric, and clinical data were collected from the EMR. This included age, sex, weight, height, body mass index (BMI), blood pressure, heart rate, diagnoses (as indicated by the ICD code), and echocardiogram reports (specifically left ventricular mass over body surface area [BSA] and left ventricular mass Z-score). If more than one blood pressure measurement was taken at a visit, we took the average of all measurements taken at the visit. We categorized patients with elevated BP into isolated systolic elevation, isolated diastolic elevation, or combined elevation. For patients with elevated blood pressure, the EMR data was reviewed to assess for appropriate management, including whether a follow-up visit was scheduled, if the patient was sent back to their family physician, if a diagnosis of white-coat hypertension was made, or if pharmacological or non-pharmacological management for hypertension was initiated. If these data were missing from the EMR, a chart review was performed. A follow-up visit was considered scheduled if it occurred within one year of the initial visit. A patient was considered lost to follow-up if a follow-up visit was scheduled but not attended. The one-year timeframe was chosen to enable a more precise analysis of blood pressure trends while ensuring standardization for comparability of the results.

### Blood pressure definitions

The blood pressure index was calculated based on systolic and diastolic blood pressure values relative to the 95th percentile, as per the 2017 AAP hypertension guidelines (28). A BP measurement was considered hypertensive when the blood pressure index exceeded 1.0, and a diagnosis of hypertension was established when this hypertensive blood pressure was measured over three consecutive visits. Isolated systolic hypertension is diagnosed when the blood pressure index exceeded 1.0 for the systolic blood pressure values only. Isolated diastolic hypertension is diagnosed when the blood pressure index exceeded 1.0 for the diastolic blood pressure values only. Combined hypertension is diagnosed when blood pressure index exceeded 1.0 for both systolic and diastolic blood pressure values. Elevated blood pressure was considered appropriately addressed if a follow-up visit was scheduled, if a diagnosis of white-coat hypertension was made, or if pharmacological or non-pharmacological management was initiated.

### Statistical Analysis

All statistical analyses were performed using R statistical programming (Version 4.2.1; (29)). Cohort characteristics were reported overall and by hypertension diagnosis as frequencies for categorical variables and as median with interquartile range (IQR) or mean with standard deviation (SD) for variables with skewed or normal distributions, respectively. Categorical variables were compared using the chi-squared test, and continuous variables were compared using the Kruskal-Wallis rank sum test. Holm’s method was used to adjust for multiple testing. BMI Z-scores were calculated based on CDC data.

## Results

During the study period, 1,469 children aged 1 to 18 years old who were newly referred to the nephrology clinic and had complete data for calculating a blood pressure (BP) index were included. At the first visit, for those patients with complete age, sex, height or blood pressure measurements, 283 patients (19.3%) were identified as having elevated BP, with 126 (8.6%) having elevated systolic blood pressure (SBP), 108 (7.4%) having elevated diastolic blood pressure (DBP), and 49 (3.3%) having both elevated (Figure 1). Demographic characteristics were compared between patients with normal BP (n = 1,186), isolated elevated SBP (n = 126), isolated elevated DBP (n = 108), and combined elevated BP (n = 49) at the first visit (Table 2). Statistically significant differences were observed among the four groups for age (*P*< 0.0001), sex (*P*=0.004), and BMI (*P*<0.0001).

**Figure 1.**
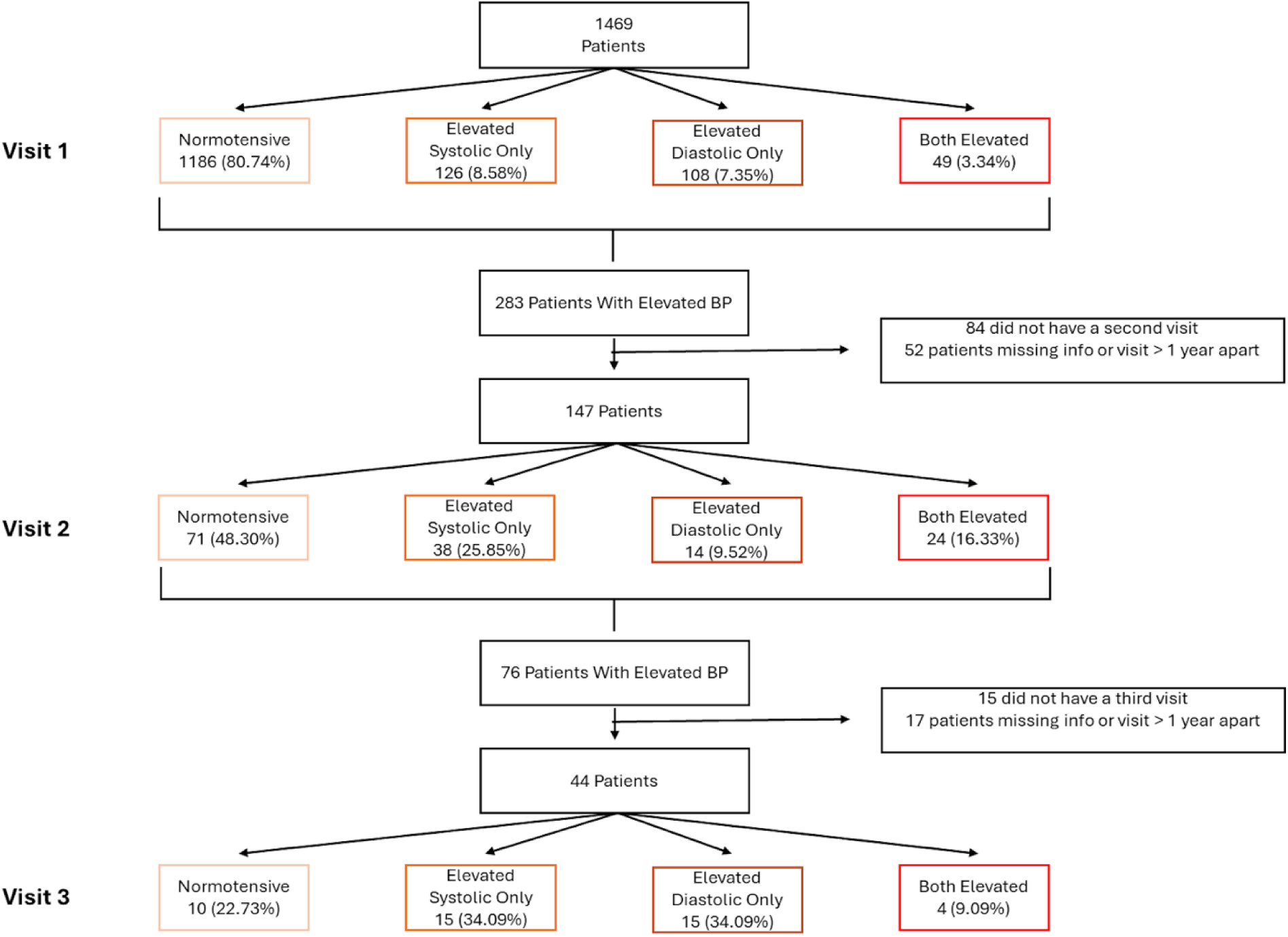
Schematic breakdown of patients that are included and excluded in the study, as well as characterization of the elevated blood pressure groups from visit 1 to visit 3.

Of the 283 patients who had an elevated BP measurement at their first visit, 136 (48.1%) were excluded after the first visit: 52 (18.4%) were found to have missing clinical data and 84 (29.7%) did not have follow-up visits (Table 1). Of the 147 patients who completed a second visit, 76 (51.7%) had an elevated BP at their second visit. Between the second and third visit, 32 patients (42.1%) were excluded: 15 (19.7%) due to missing clinical data and 17 (22.4%) due to missed follow-up visits. By the third visit, 34 of the remaining 44 patients (77.3%) were diagnosed with hypertension (HTN), including 15 with isolated systolic HTN (34.1%), 15 with isolated diastolic HTN (34.1%), and 4 with both systolic and diastolic HTN (9.1%). The median (IQR) follow up time between the 1^st^ and 2^nd^ visit for the 147 participants who had had complete info and a follow up within 12 or less months was 3.7 (1.5, 6.4) months. The median (IQR) follow up time between the 2^nd^ and 3^rd^ visit for the 44 participants who had complete information and a follow up within 12 months was 3.7 (3.0, 6.3) months.

**Table 1.** Distribution of patients with normal blood pressure and elevated blood pressure groups across the three visits, and number of patients who are excluded throughout.

|  | Visit # |  |  |
| --- | --- | --- | --- |
|  | Visit # 1 | Visit #2 | Visit #3 |
| **Number of Patients** | 1469 | 283 | 76 |
| **Number of Patients Excluded** | NA | 136 (48.06%) | 32 (42.11%) |
| Did not have record of next visit | NA | 84 (29.68%) | 15 (19.74%) |
| Missing Info or visit is greater than a year apart | NA | 52 (18.37%) | 17 (22.37%) |
| **Number of Patients After Exclusion** | 1469 | 147 | 44 |
| **Normotensive** | 1186 (80.74%) | 71 (48.30%) | 10 (22.73%) |
| **Elevated BP:** | 283 (19.26%) | 76 (51.70%) | 34 (77.27%) |
| Isolated Elevated SBP | 126 (8.58%) | 38 (25.85%) | 15 (34.09%) |
| Isolated Elevated DBP | 108 (7.35%) | 14 (9.52%) | 15 (34.09%) |
| Combined Elevated BP | 49 (3.34%) | 24 (16.33%) | 4 (9.09%) |

**Table 2.** Demographics of the patients in the nephrology clinics in their first visit as grouped into normotensive, isolated elevated systolic blood pressure, isolated elevated diastolic blood pressur or combined elevated blood pressure group.

| Characteristic | Overall<br>N = 1,469<sup>1</sup> | Normotensive N<br>= 1,186<sup>1</sup> | Systolic only<br>N = 126<sup>1</sup> | Diastolic only<br>N = 108<sup>1</sup> | Both N = 49<sup>1</sup> | p-<br>value<sup>2</sup> | q-<br>value<sup>3</sup> |
| --- | --- | --- | --- | --- | --- | --- | --- |
| Age (years) | 9.8<br>(5.2, 14.5) | 10.2<br>(5.8, 14.6) | 13.6<br>(9.4, 16.1) | 2.4<br>(1.5, 4.1) | 7.0<br>(2.5, 13.0) | <0.001 | <0.001 |
| Sex |  |  |  |  |  | 0.004 | 0.004 |
| Female | 676 (46%) | 569 (48%) | 56 (44%) | 35 (32%) | 16 (33%) |  |  |
| Male | 793 (54%) | 617 (52%) | 70 (56%) | 73 (68%) | 33 (67%) |  |  |
| Weight (kg) | 35.1<br>(19.1, 58.4) | 36.2<br>(20.5, 56.7) | 64.5<br>(35.7, 85.5) | 13.1<br>(11.2, 15.8) | 27.1<br>(16.5, 48.9) | <0.001 | <0.001 |
| (Missing) | 5 | 3 | 1 | 1 | 0 |  |  |
| Height (cm) | 138.8<br>(111.1, 162.6) | 140.6<br>(114.1, 162.8) | 158.6 (137.1, 171.2) | 89.2<br>(80.2, 101.1) | 126.4<br>(91.0, 159.7) | <0.001 | <0.001 |
| BMI (kg/m2) | 18.2<br>(16.1, 22.3) | 18.0<br>(16.1, 21.9) | 24.3<br>(20.2, 30.2) | 16.8<br>(15.5, 18.5) | 18.5<br>(16.5, 24.3) | <0.001 | <0.001 |
| (Missing) | 5 | 3 | 1 | 1 | 0 |  |  |
| BMI Z-score | 0.6<br>(-0.3, 1.4) | 0.5<br>(-0.3, 1.2) | 1.6<br>(0.7, 2.0) | 0.2<br>(-0.6, 1.0) | 1.5<br>(-0.3, 2.0) | <0.001 | <0.001 |
| (Missing) | 114 | 58 | 4 | 44 | 8 |  |  |
| Pulse Rate | 90.0<br>(78.0, 104.0) | 87.0<br>(76.0, 100.0) | 89.0<br>(76.0, 102.0) | 110.0<br>(104.0, 122.0) | 111.0<br>(100.0, 127.0) | <0.001 | <0.001 |
| (Missing) | 28 | 17 | 3 | 6 | 2 |  |  |
| Average systolic | 103.0<br>(94.0, 112.5) | 101.6<br>(93.5, 109.5) | 129.6 (121.3, 139.3) | 97.0<br>(89.3, 103.0) | 125.3<br>(110.0, 138.3) | <0.001 | <0.001 |
| Average diastolic | 60.0<br>(55.0, 66.0) | 59.0<br>(54.0, 64.0) | 67.6<br>(61.5, 73.5) | 66.0 (63.0, 73.0) | 79.8<br>(73.3, 85.3) | <0.001 | <0.001 |
| Average systolic Z-score | 0.4<br>(-0.4, 1.1) | 0.2<br>(-0.5, 0.7) | 2.2<br>(1.9, 2.6) | 0.8<br>(0.1, 1.3) | 2.6<br>(1.9, 3.2) | <0.001 | <0.001 |
| Average diastolic Z-score | 0.2<br>(-0.4, 1.0) | 0.0<br>(-0.6, 0.6) | 0.6<br>(0.1, 1.1) | 2.1<br>(1.9, 2.5) | 2.3<br>(1.9, 3.0) | <0.001 | <0.001 |

<sup>1</sup>Median (Q1, Q3); n (%)
<sup>2</sup>Kruskal-Wallis rank sum test; Pearson's Chi-squared test
<sup>3</sup>Holm correction for multiple testing

Among patients excluded due to missing follow-up information, 35.7% (30 of 84) from visit 1 had their elevated BP addressed through clinician actions, such as scheduling follow-up appointments (even if missed), diagnosing white-coat hypertension from ambulatory monitoring or initiating hypertension treatment (Table 3). At visit 2, a higher proportion (11 of 15, 73.3%) had their elevated BP addressed by clinicians. Patients whose BP was addressed at visit 1 were more likely to have appointments primarily related to blood pressure management (22 of 30, 73.3%) compared to those whose BP was not addressed (7 of 53, 13.2%) (Table 4). Additionally, patients whose BP was not addressed tended to have isolated elevated DBP (32 of 53, 59.3%) compared to only 1 patient with isolated elevated DBP (3.3%) in the addressed group.

**Table 3.** Breakdown of patients with elevated blood pressure who did not have records of their next clinic appointment.

|  | End of Visit # |  |
| --- | --- | --- |
|  | Visit #1 | Visit #2 |
| **Number of patients excluded** | 283 | 76 |
| **Number of patients did not have record of next visit** | 84 | 15 |
| **Elevated BP was addressed<sup>2</sup>** | 30 (35.71%) | 11 (73.33%) |
| Follow-up visit scheduled<sup>3</sup> | 19 | 10 |
| Lost in follow-up | 9 | 5 |
| Community Nephrology Appt | 1 | 0 |
| Tele-visit | 3 | 5 |
| Not within study period | 6 | 0 |
| Transfer back to FP | 5 | 0 |
| White-coat hypertension | 12 | 6 |
| Treatment started | 13 | 8 |
| Antihypertensive | 3 | 5 |
| Lifestyle change | 10 | 6 |
| **Elevated BP was not addressed** | 54 (64.29%) | 4 (26.67%) |

**Table 4.** Comparison of patients who had their elevated BP addressed versus those who did not.

|  | End of Visit # 1 Without Subsequent Visit<br>(n= 84) |  | End of Visit # 2 Without Subsequent Visit<br>(n= 15) |  |
| --- | --- | --- | --- | --- |
|  | Elevated BP<br>Addressed<br>(n= 30) | Elevated BP Not<br>Addressed<br>(n= 54)* | Elevated BP<br>Addressed<br>(n= 11) | Elevated BP Not<br>Addressed<br>(n= 4) |
| **Isolated Elevated SBP** | 24 (80.00%) | 10 (20.37%) | 7 (63.64%) | 1 (25.00%) |
| **Isolated Elevated DBP** | 1 (3.33%) | 32 (59.26%) | 0 (0%) | 2 (50.00%) |
| **Combined Elevated BP** | 5 (16.67%) | 11 (20.37%) | 4 (36.36%) | 1 (25.00%) |
| **Visit is related to BP** | 22 (73.33%) | 7 (12.96%) | 11 (100.00%) | 2 (50.00%) |
| **Visit is not related to BP** | 8 (26.66%) | 47 (87.04%) | 0 (0%) | 2 (50.00%) |

\* 1 patient in the elevated BP not addressed group did not have any record of nephrology clinics and therefore was not categorized into any of the 3 BP group

Patients with isolated elevated DBP at the first visit (median age: 2.4 years) and combined elevated BP (median age: 7.0 years) were younger compared to those with isolated elevated SBP (median age: 13.6 years) and normal BP (median age: 10.2 years). A higher proportion of males were observed in the isolated elevated DBP (68%) and combined elevated BP (67%) groups compared to a more equal sex distribution in the normal BP and isolated elevated SBP groups. Patients with isolated elevated SBP had a higher average BMI of 24.3 kg/m² (20.2, 30.2) compared to those with isolated elevated DBP (16.8 kg/m² (15.5, 18.5)), combined elevated BP (18.5 kg/m² (16.5, 24.3)), and normal BP (18.0 kg/m² (16.1, 21.9)). Finally, patients with isolated, elevated DBP and combined, elevated BP had higher resting heart rates (110 bpm (104, 122) and 111 bpm (100, 127), respectively) compared to those with isolated elevated SBP (89 bpm (76, 102)) and normal BP (87 bpm (76, 100)). When comparing absolute number of blood pressure measurements taken, patients with isolated elevated DBP were more likely to have only a single BP measurement recorded per clinic visit (61 of 108, 56.5%) compared to those with isolated elevated SBP (10 of 126, 7.9%) or combined elevated BP (5 of 49, 10.2%) (Figure 2).

**Figure 2.**
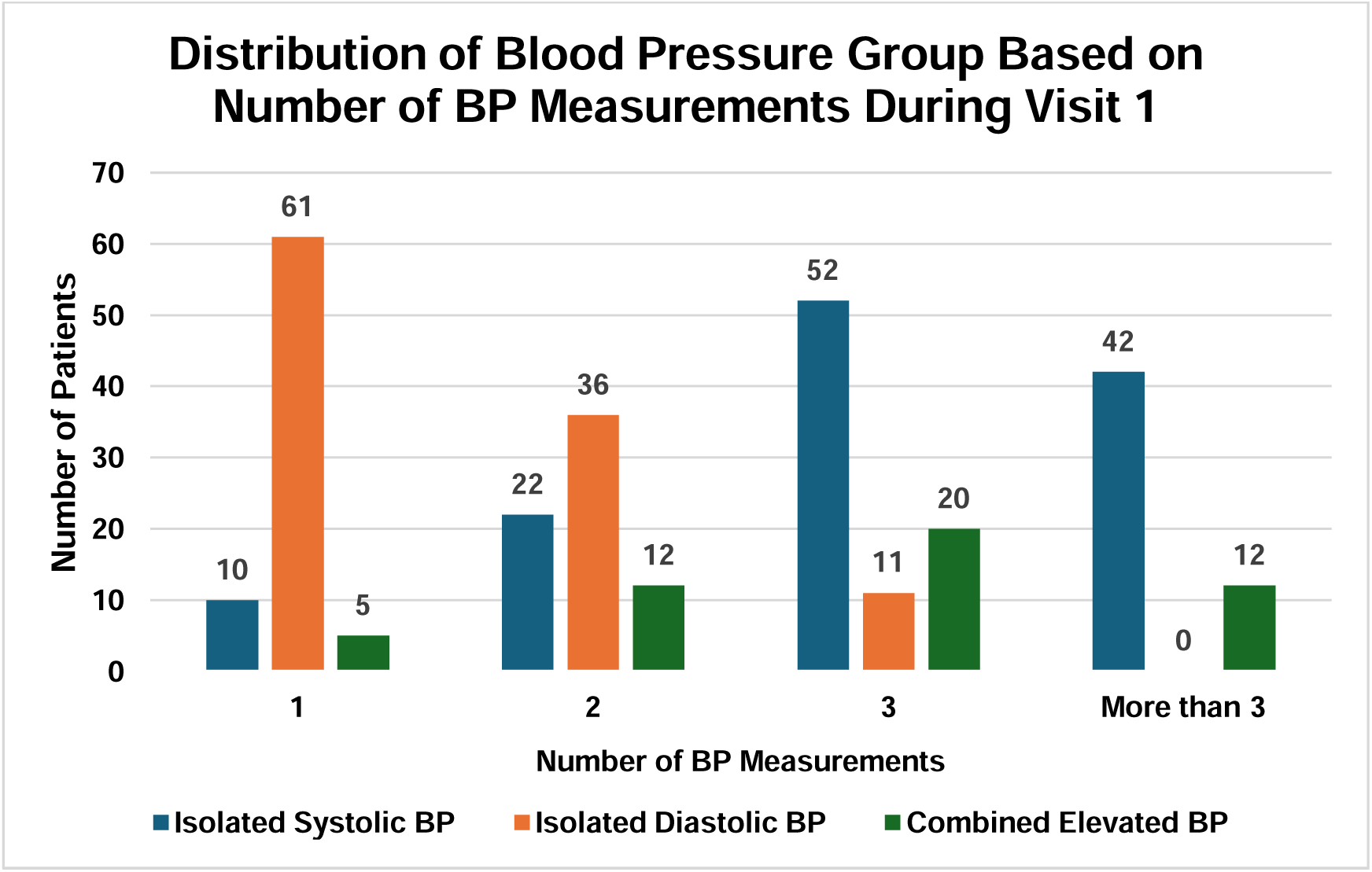
Distribution of blood pressure groups based on number of blood pressure measurements done during visit 1.

## Discussion

The prevalence of pediatric HTN is increasing, driven largely by the obesity epidemic (12). Diagnosing HTN in children and adolescents poses unique challenges due to complex diagnostic criteria that vary by age, sex, and height. This study analyzed the potential utility of extracting EMR data to improve the diagnosis and management of pediatric HTN. Patient attrition posed significant limitations to our study. Among patients with elevated BP, 136 (48.1%) and 32 (42.1%) were excluded after the first and second visits, respectively, due to missing follow-up records or clinical data as obtained using the EMR. Upon manual chart review and analysis of patients who did not have records of a follow-up appointment, 35.7% after visit 1 and 73.3% after visit 2 had their elevated BP addressed through one of the following: a scheduled follow-up that was missed, a diagnosis of white-coat hypertension, or the initiation of treatment. This highlights the critical issue of EMR data quality. Errors and fragmentation in EMRs are common, often resulting from rushed documentation, incorrect data entry, or incomplete patient histories. Data quality in EMRs has been a well-discussed topic in other studies. Specifically, the English National Health Service reported significant misclassifications in hospital admissions, including approximately 20,000 adults having been documented as being admitted to pediatric services, 17,000 males to obstetrical services, and 8,000 males to gynecology services (30). This problem was apparent in our study where many patients were excluded for missing clinical data. Furthermore, reliance on a single EMR system can obscure a patient’s complete clinical history, particularly during care transitions to other institutions or adult providers. This is evident in our study, where some patients’ follow-up care occurred in the community, or they had transitioned to adult nephrology care. By highlighting the challenges in maintaining accurate and comprehensive EMR data, this creates an opportunity to evaluate ways to improve our EMR data quality. For example, training for healthcare professionals exists, focusing on accurate data entry or even promoting interoperability between different EMR systems to allow for more comprehensive data collection. In addition, EMRs can be built to identify outlier data whereby automatic notification prompts would notify clinicians to repeat BP measurements that exceed the 90th percentile, for example.

Our study also revealed gaps in the management of elevated BP, particularly when the primary referral is unrelated to HTN or the clinical presentation is isolated diastolic hypertension (IDH). Notably, patients whose elevated BP was not addressed were more likely to have isolated elevated DBP or visits unrelated to BP management. When a patient presented with isolated diastolic blood pressure elevations, they typically had only a single BP measurement taken (56.5%), compared to the multiple measurements taken for patients with isolated or mixed elevated SBP (7.9% and 10.2%). Translated clinically, this indicates that elevated DBP measurements were more likely to be overlooked, potentially reflecting the perceived benign nature of this phenomenon. Underdiagnosis of hypertension is a known issue for both the pediatric and adult population (31–33). In a cross-sectional retrospective study, Moin *et al*. found that only 14.6% and 41.8% of children meeting criteria for prehypertension and hypertension, respectively, were diagnosed in their EMRs (33). Contributing factors include the complexity of diagnostic criteria (34), inadequate staff training (35), and challenges in obtaining accurate BP measurements (36).

This study has several limitations. As a retrospective analysis based on single center EMR data, our findings are subject to selection bias and may not be generalizable to the broader population. Missing and incomplete data led to the exclusion of a substantial portion of our cohort. Additionally, the study period overlapped with the COVID-19 pandemic, likely contributing to follow-up challenges. Finally, the small sample size in subsequent visits limited our ability to conduct more comprehensive analyses. Strengths of this study included our supplementation of EMR data with a manual review of clinical charts to confirm reasons for missing EMR data. In addition, the categorization of patients into different hypertension phenotypes (ISH, IDH and combined hypertension) allowed for a more in-depth analysis and further revealed that elevated DBP readings may be discarded. This is critical, as recent work from our group revealed that those with IDH had similar risk factors for LVH (37).

## Conclusion

This study identifies some of the challenges associated with studying pediatric hypertension using an EMR, particularly highlighting missing values and insufficient measurements as problematic. EMR data quality is paramount. Future studies seeking to optimize clinical procedures with subsequent reassessment of EMR utility are under consideration.

## Ethical Approval

Approved by REB (22/128X) at The Children’s Hospital of Eastern Ontario, Ottawa, ON, Canada

## Consent to Participate

Not applicable as it is a retrospective chart review

## Data availability

All data produced in the present work are contained in the manuscript.

